# Multivariate brain–cognition covariance supports the criterion validity of cognitive screening performance

**DOI:** 10.64898/2026.02.26.26347152

**Authors:** Kristīne Šneidere, Nauris Zdanovskis, Zane Anna Litauniece, Agnese Ušacka, Anita Ilze Gulbe, Zigmunds Freibergs, Ainārs Stepens, Kristīne Mārtinsone

## Abstract

There is a predicted increase in older adults presenting with mild to severe cognitive impairment. Screening tools with high sensitivity are the first frontier in identifying a cognitive pathology; however, to ensure that they are measuring the intended concept or criterion, thorough psychometric procedures should be followed. In this study, convergent criterion validity of Riga Cognitive Screening Task was measured, using cortical thickness of regions of interest as the criterion. 106 older adults (*M_age_* = 70.49, SD =8.08, 35.8% male) with varying levels of cognitive functioning were involved in the study. All participants underwent cognitive assessment with the screening task and a 3T MRI. Cortical thickness of selected temporal and parietal regions was used as a brain measure. Behavioural Partial Least Squares Correlation was conducted and one latent variable was extracted. The results confirmed that Riga Cognitive Screening Task shows good criterion validity, suggesting successful use for screening.

## Introduction

A recent study using the data from the Survey of Health, Ageing and Retirement in Europe (SHARE) came to a striking conclusion, suggesting that the actual prevalence of dementia in Europe varies from 5 – 22% across countries(1), contradicting the current statistics of 2.02% of the population. This discrepancy points to widespread underdiagnosis. Dementia comes with a wide variety of burdens. Economical cost of dementia in European countries varies from 7938 – 73 712 EUR yearly, with the cost increasing with the severity of the disease(2), pointing to significant expenses to the health care system and individuals.

Early detection of symptoms associated with pathological cognitive decline is critical, as it allows for timely intervention, thus prolonging the quality of life(3). Here cognitive screening often serves as the first diagnostic step and plays a vital role in primary, secondary and tertiary prevention, by facilitating early detection, as well as diagnostic accuracy(4,5). Nevertheless, while currently there is a wide variety of used tools, such as Mini-Mental State Exam (MMSE), many of those show bias towards education and culture(6), as well as overlook early deficits in reasoning, semantic processing, executive control or pragmatic reasoning(7). With this in mind, Riga Cognitive Screening Task (RiTa) was developed, integrating information on lifestyle and life-experiences (i.e. cognitive reserve) with cognitive assessment scales, aiming to obtain an integrative profile of cognitive performance in older adults with suspected mild cognitive impairment or other forms of pathological cognitive decline(8).

The content of a screening test, particularly considering the linguistic and cultural aspect, can significantly affect the perception of the test questions and tasks, thus also having an impact on the results. Therefore, a test or a task that has only been developed without applying the proper psychometric procedures could yield higher rates of false-positive or false-negative scores (9), thus contributing to misdiagnosis. A psychological test is essentially a systematic procedure developed to measure a behaviour (e.g. cognitive functioning) in a planned, uniform and thorough way, allowing to administer the test and evaluate the behaviour equally across patients or participants (10). The main criteria involved in the development and general assessment of the psychometric properties of RiTa have been discussed in the publication by Šneidere et al.(8); however, a significant psychometric indicator is criterion validity that has been discussed through the total scores in the aforementioned article. Criterion validity is assessed to determine the relationship between the newly developed test and a specific criterion(11). Within this study, concurrent criterion validity was assessed, namely, to what extent individual differences in RiTa performance covary with cortical thickness in regions known to be vulnerable in pathological cognitive ageing.

There is a substantial body of literature suggesting an increased vulnerability of specific cortical regions in pathological cognitive ageing, with Alzheimer’s Disease (AD) providing one of the most extensively characterised models of this process. Across neurodegenerative conditions, medial temporal cortex (MTL) has been consistently identified as among the earliest and most affected regions. This has been especially significant in areas like hippocampus and entorhinal cortex, with entorhinal cortex exhibiting the earliest and most severe neuronal loss, potentially due to vulnerability to Tau pathology (12,13). Parahippocampal gyrus has also been found to be affected at early stages, with both – cortical atrophy and disrupted connectivity contributing to memory impairment (14,15).

While MTL structures are often considered sensitive early biomarkers of pathological ageing, lateral temporal cortex structures have also shown early and pronounced cortical thinning. The fusiform gyrus (FG), a structure involved in facial recognition, object processing, and multisensory integration (16–18), supports higher-order perceptual and social cognitive functions. Post-mortem, region-specific epigenomic and transcriptomic studies in AD have demonstrated that the FG shows distinct molecular profiles compared to other cortical regions, suggesting differential vulnerability that may track disease progression beyond traditional neuropathological staging schemes(19). Similarly, the inferior temporal gyrus has been found to contribute to cognitive and behavioural functions associated with social interaction, e.g. visual object and language comprehensions and lexical and phonologic decisions(20). In pathological ageing, volume loss in the right rostral Brodmann area 20 in the ITG has been associated with cognitive impairment and differentiating AD from late mild cognitive impairment (21). The middle temporal gyrus has likewise been shown to exhibit early vascular dysfunction, structural atrophy, and cell-type-specific transcriptomic alterations (22–24), alongside changes in semantic processing and language functions (25,26).

Beyond temporal regions, parietal cortices have been linked to more rapid clinical progression and multidomain cognitive decline (27,28). The precuneus functions as a highly connected (rich-club) hub within the default mode network (DMN)(29), integrating multimodal information while supporting visuospatial processing and episodic memory (30). The inferior parietal lobule (IPL) functions as another key neural structure that is involved in diverse cognitive roles, including attention and control in sensorimotor networks, self-referential, memory and social processes, also showing strong hemispheric specialization whereby left IPL is more significantly involved in language and semantics and the right IPL is contributing to attention orienting and social evaluation (31). Consistent reductions in cortical thickness within inferior parietal and temporal regions have been observed in Alzheimer’s disease (32). The supramarginal gyrus, in turn, is primarily associated with phonological working memory and early semantic processing (33).

Importantly, neurodegeneration associated with pathological ageing follows a spatial–temporal trajectory rather than a uniform pattern. Neuropathological and neuroimaging studies indicate that cortical thinning typically emerges first in medial temporal structures, followed by lateral temporal and parietal cortices (34). This progression implies that individuals at any given point along the ageing–pathology continuum may exhibit partially overlapping but non-identical anatomical profiles. Consequently, variability across distributed cortical regions should not be treated as noise, but rather as a meaningful signal reflecting system-level reorganization during cognitive decline.

Multivariate approaches, such as Partial Least Squares Correlation are well suited to this problem, as they identify latent patterns of covariation between distributed neural architecture and cognitive performance, without imposing *a priori* assumptions about one-to-one structure–function relationships. In this context, criterion validity can be examined at the level of coherent brain–cognition patterns rather than isolated structure–function pairs.

The aim of this study was to assess the criterion validity of the Riga Cognitive Screening Task Cognitive assessment scale using subscales of RiTa as proxies for cognitive processes and the thickness of regions from medial temporal, lateral temporal and parietal cortices as markers of brain atrophy. We hypothesised that Behavioural PLSC will identify a significant latent variable linking RiTa subscales to a distributed cortical thickness pattern, showing that lower performance across RiTa domains would also be associated with reduced thickness in temporal and parietal cortices.

## Materials and Methods

### Sample and procedure

106 older adults (*M_age_* = 70.49, SD =8.08, 35.8% male) were involved in the study. Only older native Latvian speakers with no self-reported ongoing oncological, psychiatric or neurological treatment other than cognitive impairment treatment associated with mild cognitive impairment (MCI) or phenotypically consistent with Alzheimer’s disease–type dementia were eligible for the study. Recruitment was conducted through senior organisations and clinics. To ensure eligibility, all participants underwent a short clinical interview over the phone, conducted by a neurologist. At the next step, each participant was completed the Montreal Cognitive Assessment scale (35) (MoCA) as a measure of general cognitive functioning.

Participants presented with varied levels of education; however, the majority had obtained at least high school diploma (*M_years_* = 14.01, *SD*=3.51, ranging 0 – 23). MoCA values ranged from 10 – 29 (*M*=22.27, *SD*=4.41). Within this step of the validation, all participants were included in the data analysis, independent of their MoCA score.

All participants signed written informed consent and approval was obtained from Riga Stradiņš University Ethics Committee. All data were obtained from October 3^rd^ 2024 to June 10^th^ 2025.

### Cognitive assessment

All participants underwent Riga Cognitive Screening Task (RiTa). This screening test consists of two main scales that include factors associated with prevention and prophylaxis of neurodegenerative disease, and a cognitive assessment scale, that includes 11 tasks, assessing episodic and semantic verbal memory, working memory, orientation in time and space, phonemic and semantic fluency, visuospatial abilities, attention, reasoning, inhibition and motor abilities. The pilot study of the screening tool showed good internal consistency in the Cognitive assessment scale, good differentiation between the diagnostic groups, excellent convergent validity with MoCA test and also moderate to excellent sensitivity and specificity. Within the PLS-C, results of each individual task representing a specific cognitive function from the Cognitive assessment scale, were entered in the model. Descriptive statistics for the cognitive tasks included in the PLS-C analysis are presented in the Results section (Table 1).

**Table 1.**
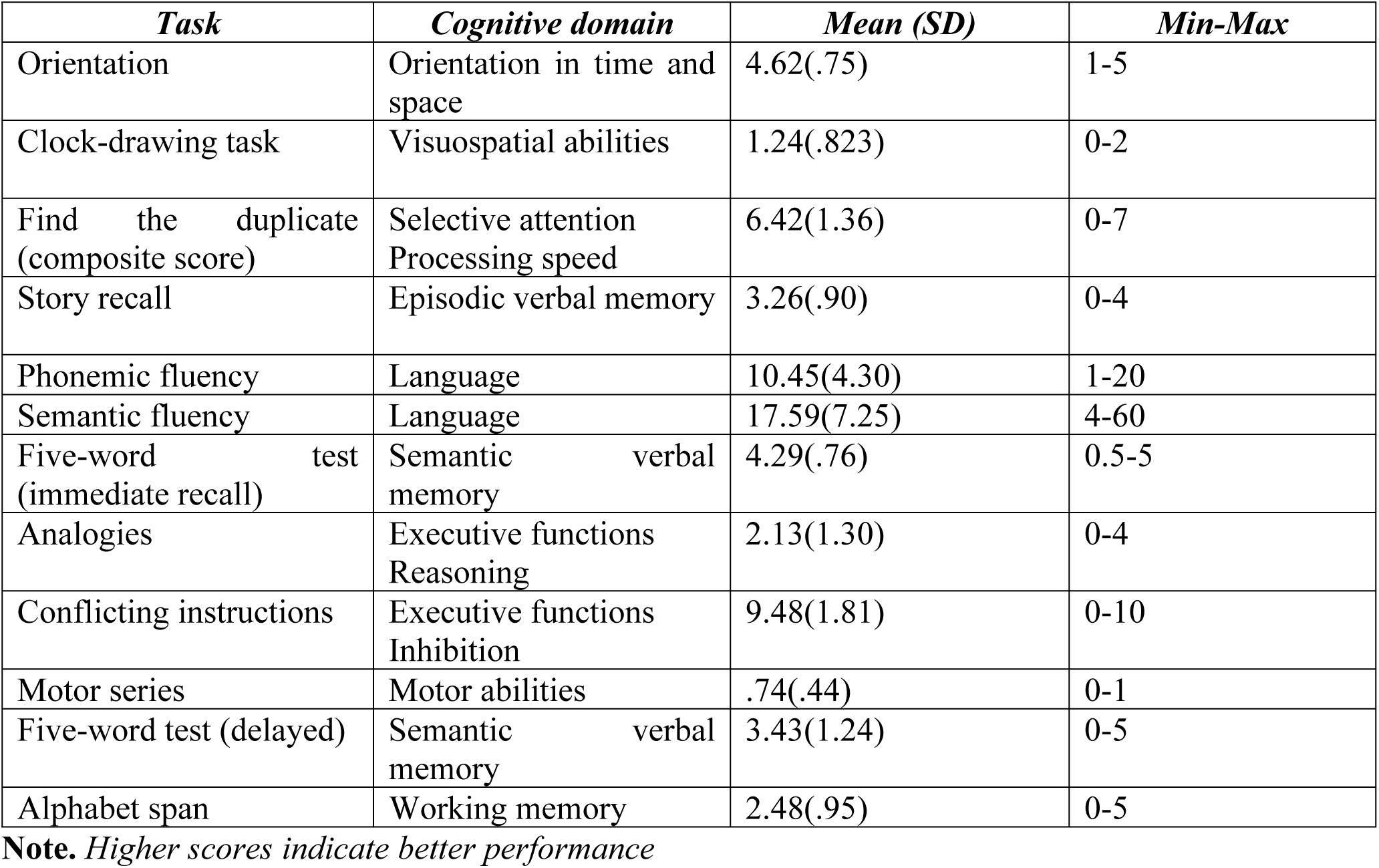
Descriptive statistics of RiTa cognitive tasks included in PLS-C analysis.

| *Task* | *Cognitive domain* | *Mean (SD)* | *Min-Max* |
| --- | --- | --- | --- |
| Orientation | Orientation in time and space | 4.62(.75) | 1-5 |
| Clock-drawing task | Visuospatial abilities | 1.24(.823) | 0-2 |
| Find the duplicate (composite score) | Selective attention<br>Processing speed | 6.42(1.36) | 0-7 |
| Story recall | Episodic verbal memory | 3.26(.90) | 0-4 |
| Phonemic fluency | Language | 10.45(4.30) | 1-20 |
| Semantic fluency | Language | 17.59(7.25) | 4-60 |
| Five-word test (immediate recall) | Semantic verbal memory | 4.29(.76) | 0.5-5 |
| Analogies | Executive functions<br>Reasoning | 2.13(1.30) | 0-4 |
| Conflicting instructions | Executive functions<br>Inhibition | 9.48(1.81) | 0-10 |
| Motor series | Motor abilities | .74(.44) | 0-1 |
| Five-word test (delayed) | Semantic verbal memory | 3.43(1.24) | 0-5 |
| Alphabet span | Working memory | 2.48(.95) | 0-5 |

**Note.** Higher scores indicate better performance

### MRI acquisition and preprocessing

All participants underwent magnetic resonance imaging (MRI) using a 3.0 Tesla scanner (General Electric (GE), Boston, MA, USA) with sequences, including 3D T1 SPGR, 3D FLAIR, High-resolution T2 hippocampal sequence, Diffusion tensor imaging (DTI) (with 32 directions, 2 b values (0 and 1000 s/mm2), and susceptibility weighted imaging (SWI). Within this study, only T1 sequence were analysed, while the other sequences were acquired as part of a broader protocol.

Structural brain segmentation was performed on T1-weighted images acquired at 3T using the standard FreeSurfer processing pipeline (recon-all, version 8.1.0.), which incorporates SynthSeg for robust whole-brain segmentation. SynthSeg is a deep learning–based approach that enables accurate segmentation across varying image contrasts and resolutions without the need for additional training or contrast-specific optimization. Within the recon-all workflow, SynthSeg was used with default parameters to generate volumetric segmentations and regional volume estimates, with internal resampling to 1 mm isotropic resolution. All segmentations were visually inspected for gross errors, and derived volumetric measures were used for subsequent analyses(36,37).

### Partial Least Squares Correlation

Behavioural Partial Least Squares Correlation (PLS-C), analysis that is specifically common in brain and behaviour design, was used (38,39). This is a symmetric multivariate technique that aims to examine relationships between pairs of data tables by identifying latent variables, that in turn maximize the shared covariance between them. In this framework, both tables are treated symmetrically, and the contribution of individual variables to each latent variable is quantified by their saliences, which can be interpreted analogously to loadings in principal component analysis(40).

#### Component selection and interpretation

Latent components were ordered according to the amount of shared covariance explained, with the first component representing the strongest multivariate brain–cognition association (see Fig 1). Components were considered for interpretation based on the magnitude of the latent correlation and the coherence and interpretability of the associated variable patterns.

**Fig 1.**
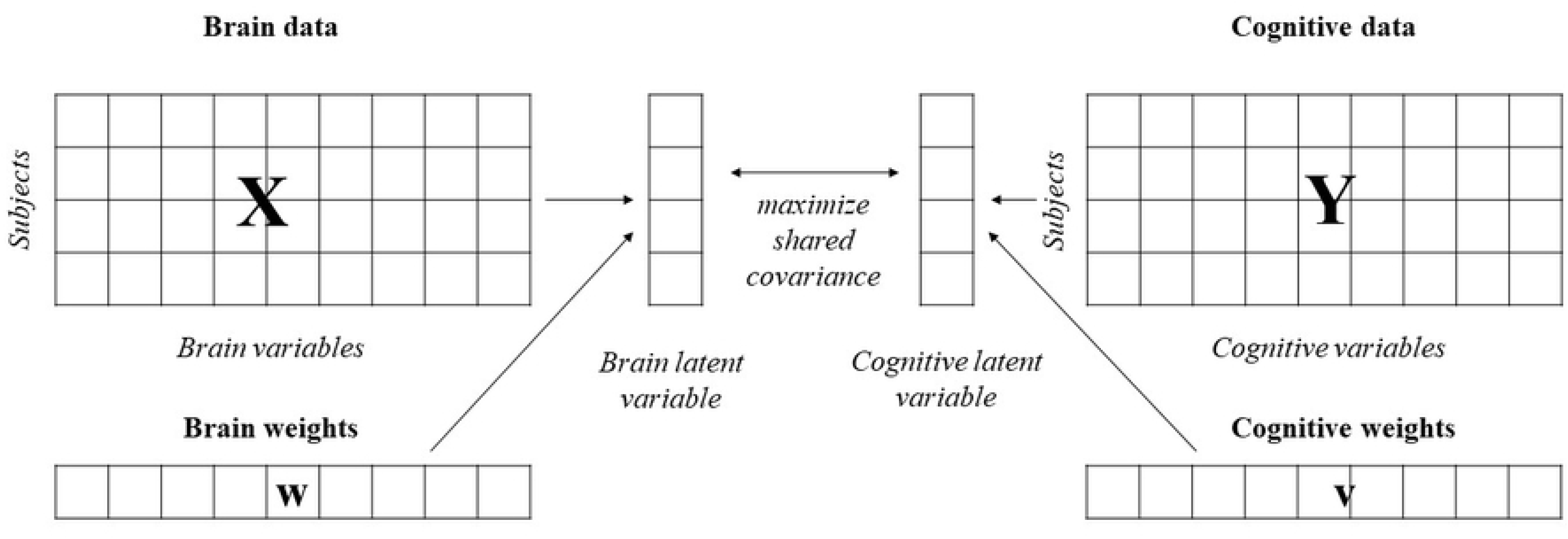
Conceptual illustration of the PLS-C framework. Multivariate brain data (X) and cognitive data (Y) are projected onto latent variables via weighted linear combinations of their respective variables (w and v).

The contribution of individual brain regions and cognitive measures to each component was assessed using saliences, which represent the weights of the original variables in the construction of the latent variables. Variables with larger absolute salience values were interpreted as contributing more strongly to the corresponding latent component. Interpretation focused on overall patterns across regions and cognitive domains rather than on individual variables in isolation.

#### Statistical considerations

Given the theory-driven selection of ROIs and the focus on identifying system-level brain–cognition patterns rather than precise ranking of individual variables, results were interpreted primarily based on the magnitude and coherence of the latent associations. The primary outcome of interest was the strength of the latent brain–cognition correlation for each component. Secondary components were interpreted cautiously and only when they represented distinct and theoretically meaningful patterns. Permutation p-values were estimated as the proportion of permuted correlations with |r| ≥ |r_obs|. To ensure the anatomical specificity of the identified multivariate thickness-cognition pattern, a theoretically unrelated control region (rostral anterior cingulate cortex) was included as a negative control region. All analyses were conducted in R using the mixOmics package(41).

## Results

### Descriptive statistics

Descriptive statistics for all cognitive tasks included in the PLS-C analysis are presented in Table 1, showing adequate variability across measures.

### Cognitive measures

Cognitive domains showed moderate to strong intercorrelations after FDR correction (Fig 2), indicating shared but non-redundant variance across domains (r= [.19,.65]). This pattern supports the use of a multivariate approach to examine brain–cognition covariance.

**Fig 2.**
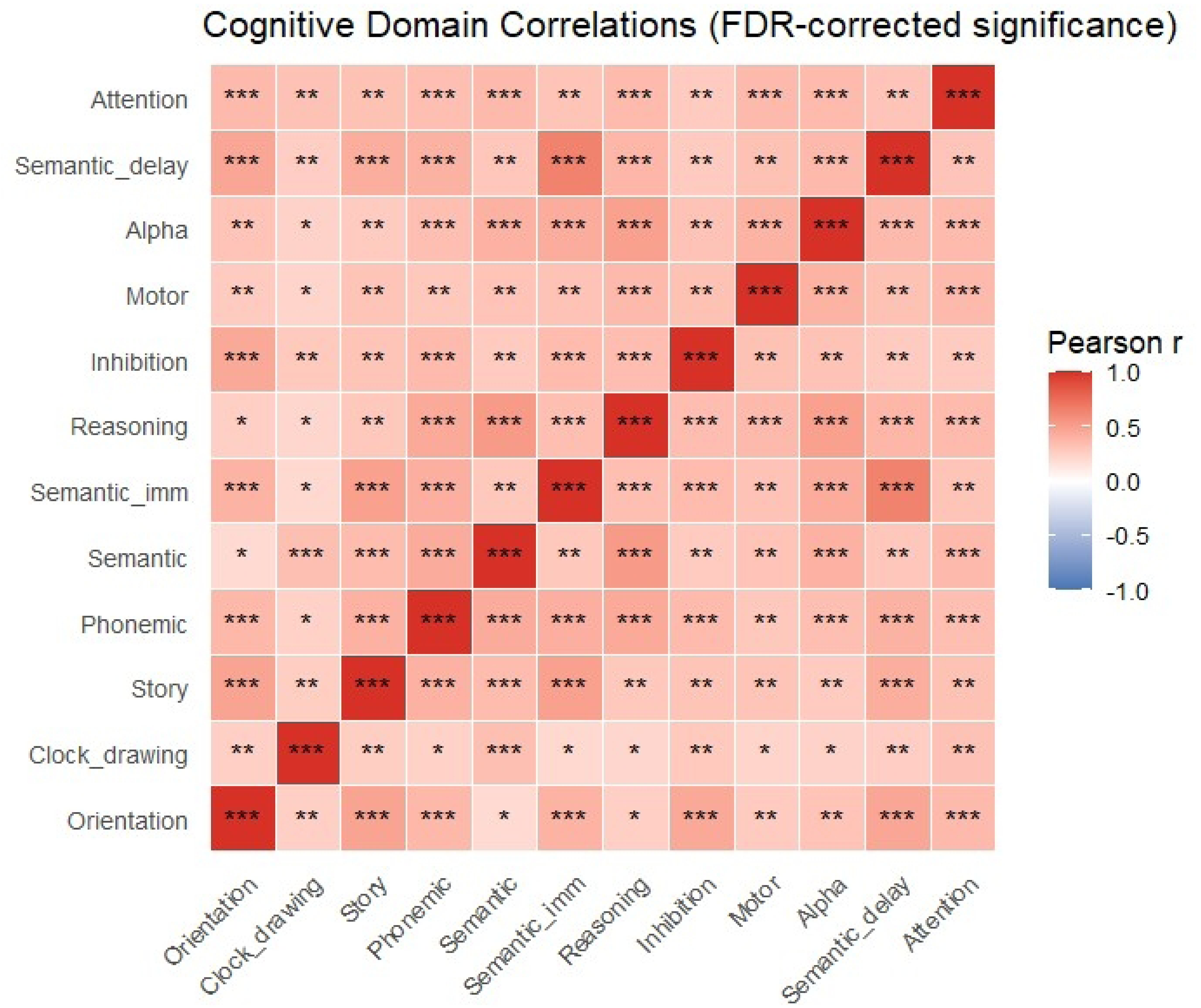
Cognitive domain correlations (FDR-corrected significance)

### Cortical thickness and cognitive dimensions

Partial least squares correlation was used to determine the relationship between the latent variables. In the X matrix, cortical thickness in selected ROIs were marked (k = 18), while the Y matrix had RiTa cognitive tasks (k = 12). All variables were standardized using z-scores to ensure the comparability of the variables. Three latent variables were extracted.

### Latent variables

The relationship between the cortical thickness in ROIs and cognitive tasks in RiTa test was captured in all three of the components. The first latent variable (LV1) showed a strong thickness-cognition association (r=.605) and was significant under permutation testing (p < .001; 1000 permutations), while second latent variable (LV2) and third latent variable (LV3) showed lower, but still strong correlations (r = .443 and r = .433, accordingly), but were not significant under permutation testing (p > .05; 1000 permutations), thus not included in further discussion.

On the cortical thickness side, largest absolute saliences were observed in 10 ROIs (w≥-.25). These regions included right hemisphere supramarginal cortex, middle temporal cortex, precuneus and inferior parietal lobule, as well as left hemisphere entorhinal cortex, middle temporal cortex, inferior parietal lobule, supramarginal cortex, precuneus and parahippocampal cortex, constituting for the core contributors to the LV1 cortical thickness pattern. In contrast, rostral anterior cingulate cortex showed negligible salience for LV1 bilaterally (w_lh_ = -.08, w_rh_ = .01) (see Fig 3).

**Fig 3.**
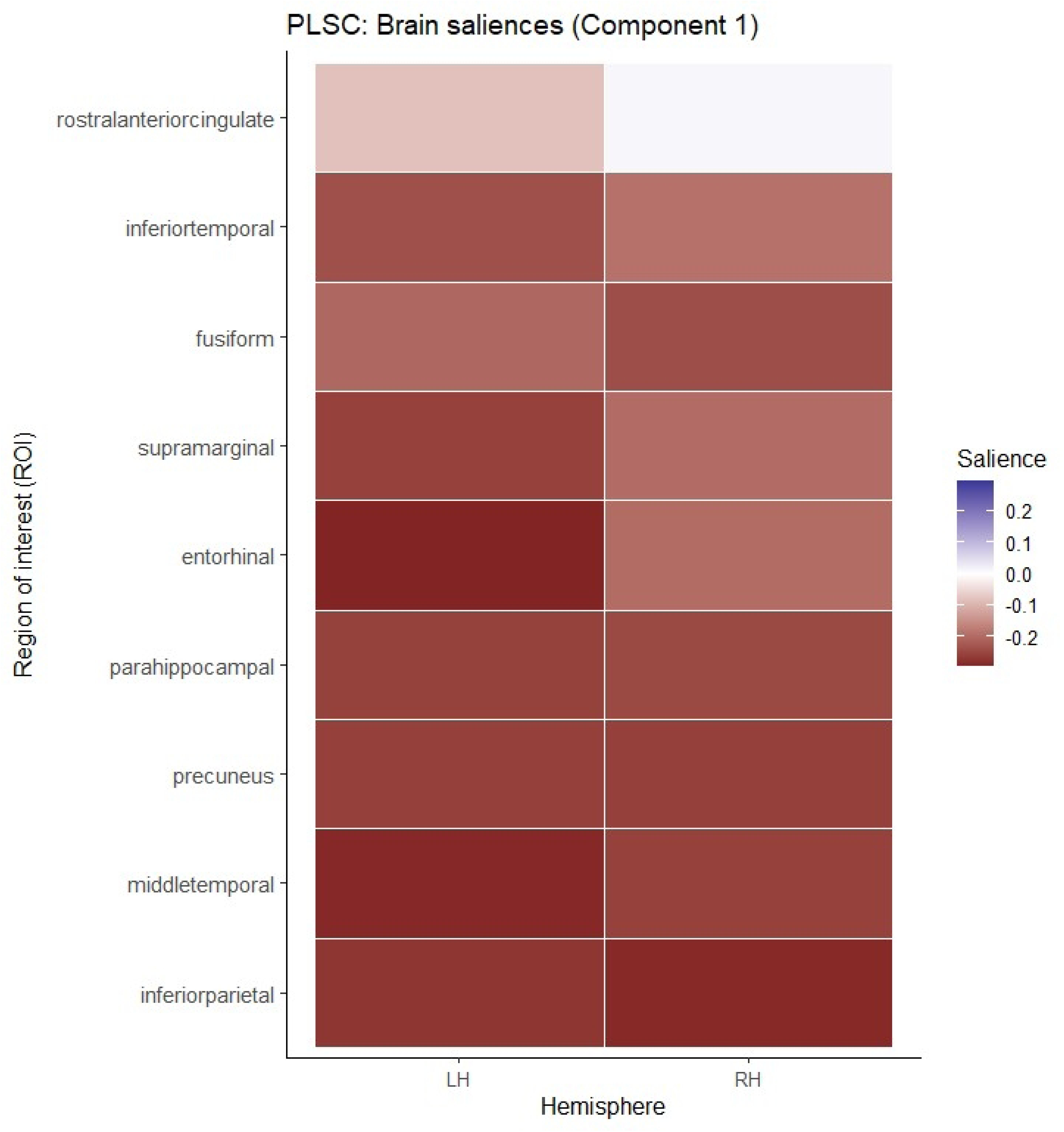
Cortical thickness saliences (LV1)

On the cognitive side, the core was made up by cognitive domains (v≥-.25), namely, inhibition, reasoning, phonemic fluency, semantic fluency, immediate and delayed semantic verbal memory, orientation in time and space and working memory. Visuospatial abilities, episodic verbal memory and selective attention and processing speed were supporting the core domains (v=-.23;-.24), while motor functions weakly contributed to the LV1 (v=-.16) (see Fig 4).

**Fig 4.**
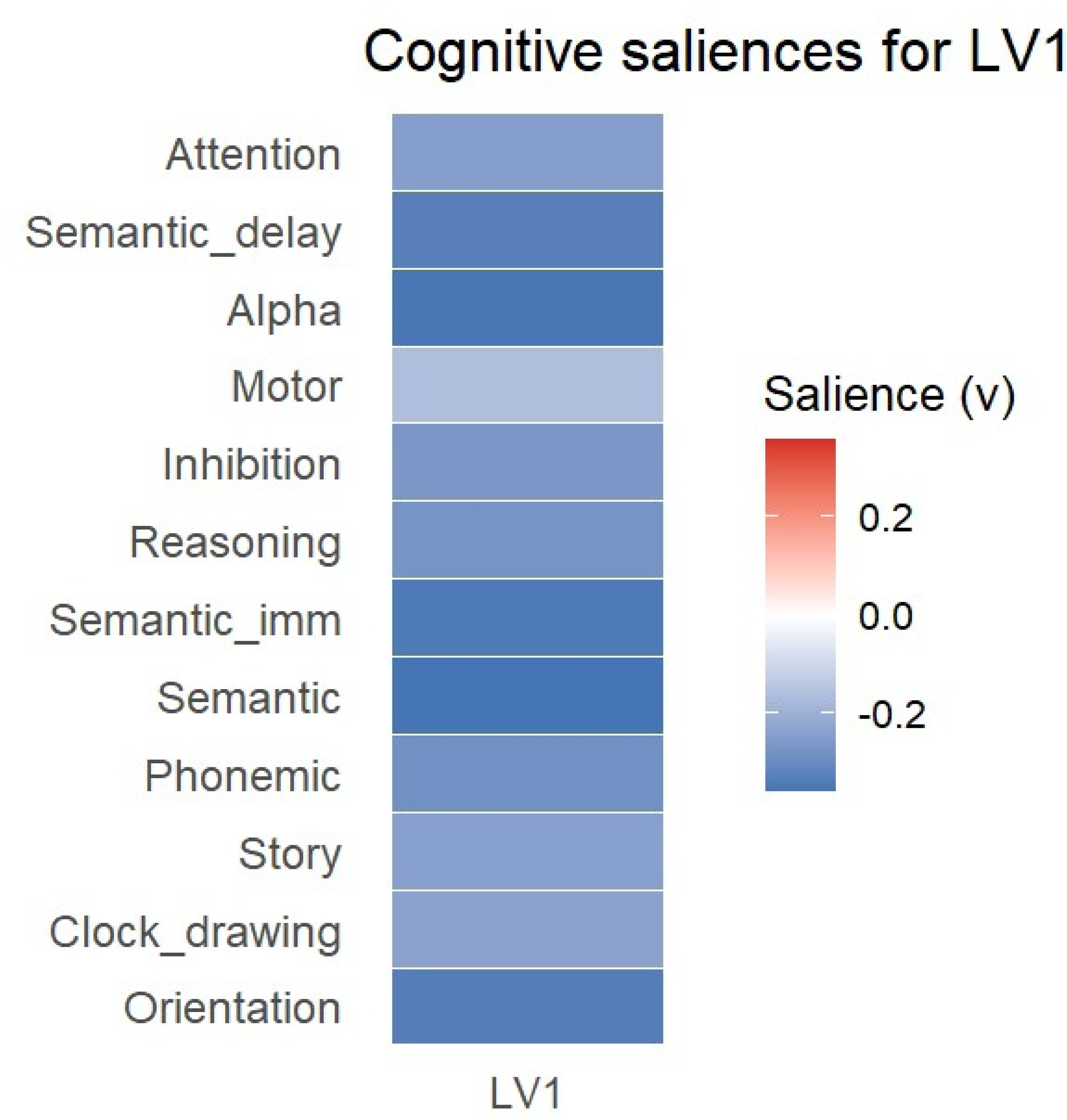
Cognitive saliences for LV1.

### Controlling for age and education

After controlling for age and education, the association between the brain thickness and cognitive scores weakened, while remaining statistically significant (r=.469, 001; 1000 permutations, p = .008) (see Fig 5). Salience similarity (baseline vs. controlled) was strong (r = .89), while Jaccard index indicated moderate reproducibility (median J =.50), overall suggesting a multivariate pattern of brain-behaviour covariation (see Fig 6).

**Fig 5.**
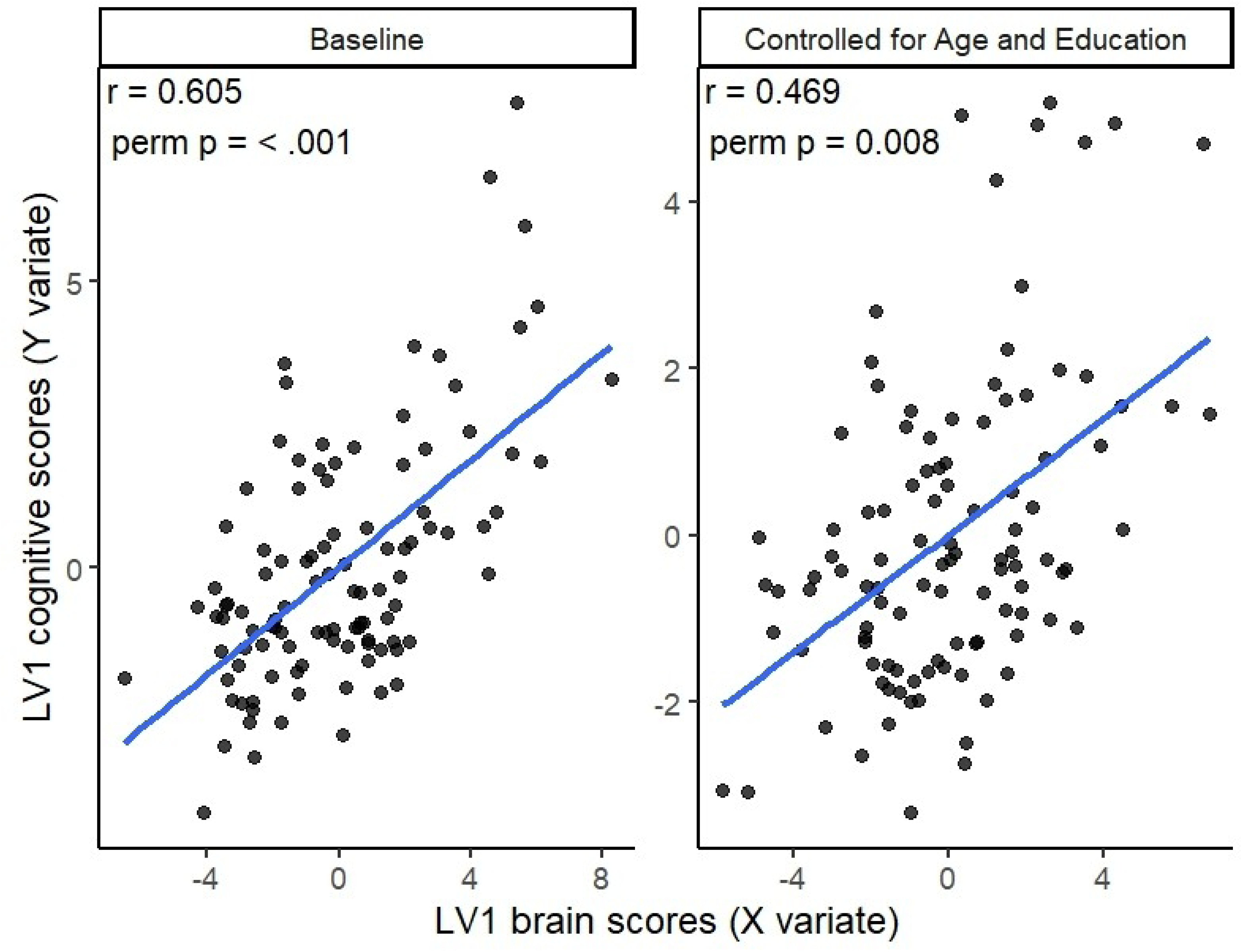
Association between the brain thickness and cognitive scores at baseline and after controlling for age and education. **Note.** Scatterplots show the relationship between LV1 brain scores (X variate) and LV1 cognitive scores (Y variate) derived from the behavioural PLS-C analysis. The left panel depicts the baseline model, while the right panel shows the model after residualising both datasets for age and education. Solid lines indicate least-squares regression fits. Permutation-based correlation coefficients and p-values are shown for each model.

**Fig 6.**
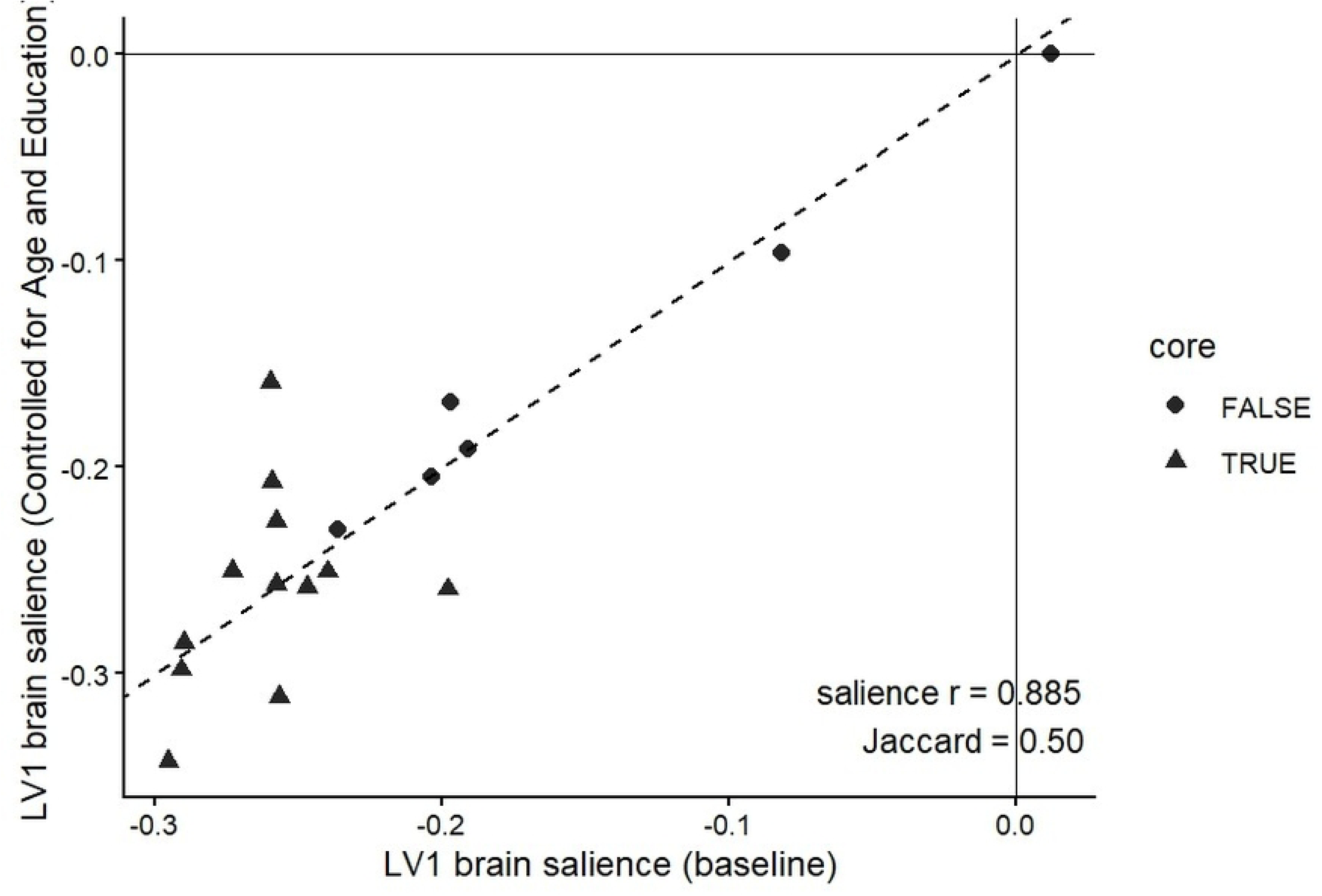
Stability of LV1 brain saliences after controlling for age and education. **Note.** Scatterplot showing the relationship between baseline LV1 brain saliences and saliences obtained after residualising for age and education. Dashed line represents the line of best fit. Triangles indicate regions classified as core contributors based on the predefined salience threshold. High salience similarity (r = .885) and moderate Jaccard overlap (J = .50) indicate preservation of the multivariate brain–cognition pattern after covariate adjustment.

## Discussion

The aim of the study was to assess the criterion validity of the Riga Cognitive Screening Task (RiTa), using behavioural partial least square correlation, expecting to identify a significant latent variable that would link together RiTa cognitive subscales with cortical thickness areas, associated with Alzheimer’s Disease. After permutation testing, we identified one meaningful latent variable (LV1) that remained significant after controlling for age and education, though dropped slightly, suggesting that age and education could contribute to the association, but does not explain the pattern.

Although additional latent variables showed moderate brain–cognition correlations, these components did not survive permutation testing and were therefore not interpreted. This pattern suggests that while multiple dimensions of covariance may exist, only the first latent variable reflects a robust and reproducible association in the present sample.

At baseline, cognitive measures indicated a broad cognitive pattern encompassing orientation in time and space, phonemic and semantic fluency, abstract reasoning, inhibition, working memory, and both – immediate and delayed recall. After controlling for age and education, the profile became more focused and excluded abstract reasoning and inhibition, instead suggesting a pattern that integrates memory and memory-related executive control. Previous studies have shown the significant impact of education on abstract reasoning, especially in older adults(42), suggesting a compensatory role of education-based cognitive reserve. Similarly, the effect of age on cognition has been observed in studies, showing higher error rates and longer response times when compared to younger adults(43), though the direct evidence of impact of education is rather sparse.

The convergence of the remaining tasks suggest that the cognitive measures do capture a shared performance with neuroanatomical measures, namely, regional cortical thickness. While the development of RiTa was focused on tasks known to being sensitive to cognitive decline, the regions of interest were chosen based on the vulnerability to age related decline, thus the regions of interest were based on previous studies, mostly exploring Alzheimer’s Disease. In this study, associated cortical thickness pattern was dominated by medial temporal, lateral temporal, and inferior parietal regions, including entorhinal, parahippocampal, fusiform, and temporal cortices. These regions are commonly implicated in memory representation(44), contextual integration, and associative processing(45), thus overall supporting the memory representation and integration processes known to underlie performance on the cognitive tasks.

While the ROI was mostly focused on the cortical regions vulnerable to Alzheimer’s Disease, we included a variety of participants that presented with extensively varied cognition (MoCA scores ranging from 10 – 30). While potentially controversial, this approach likely enhanced variance along memory-related dimensions and cut across categorical diagnoses that might increase criterion contamination. Two well-known limitation in concurrent criterion validity are the use of clinical diagnoses, as it might prioritize symptom description over the etiology and context of the disease, and reliance on criterion characteristics at a single point of measure, ignoring the continuum of the actual symptoms(46). Bypassing the division in clinical vs. control group allows to evaluate whether cognitive variability aligns with the biological variation in the cortical structures.

Overall, these findings demonstrate that criterion validity can be meaningfully assessed using multivariate brain-behaviour correspondence rather than isolated correlations. Such an approach is particularly relevant for cognitive screening tools, where performance reflects distributed systems rather than discrete cognitive modules.

A major limitation of the study is that the criterion validity was assessed cross-sectionally, thus mitigating the predictive value of the results. Furthermore, cognitive data obtained at one point in time can be impacted by external factors, such as sleep deprivation, lack of motivation or tiredness, thus challenging the objectivity of cognitive measures. Additionally, the use of cortical thickness as the sole criterion limits conclusions about the neural mechanisms related to cognitive performance. Lastly, the mean years of education indicate that most of the participants had at least high school education, that can have impact on the overall cognition; nevertheless, this is historically country specific.

## Conclusions

This study was set out to establish the criterion validity of Riga Cognitive Screening Task (RiTa), combining cognitive assessment and cortical thickness data. These results confirmed that RiTa cognitive assessment scale shows good criterion validity, suggesting that the screening task could be used for screening for cognitive dysfunction, especially when considering memory-associated impairments. This research has demonstrated that PLS-C could be used as a valuable method in testing criterion validity.

## Data Availability

Data are avaliable from Riga Stradiņš University Institutional Data Access after the embargo period.

## Acknowledgements

This research is conducted under the Postdoctoral “Grant “Development and validation of Riga Cognitive Screening Task (RiTa)” (RSU-PG-2024/1-0011), as part of the project “RSU Internal and RSU with LASE External Consolidation” (Project No. 5.2.1.1.i.0/2/24/I/CFLA/005), funded by the European Union Recovery and Resilience Facility and the budget of the Republic of Latvia.

